# Archetype-Based Stratification to Inform Malaria Intervention Targeting in Zambia

**DOI:** 10.1101/2025.08.13.25333585

**Authors:** Chilochibi Chiziba, Japhet Chiwaula, Busiku Hamainza, Sampa Chitambala-Otiono, Sheetal Silal

## Abstract

Although Zambia does not rank among the highest contributors to global malaria cases, in 2023, it had approximately eleven million cases. Furthermore, Zambia’s malaria incidence distribution is spatially heterogeneous at fine scales such as health facility catchment areas (HFCAs). HFCA-level malaria incidence stratification currently guides implementation and targeting approaches. We aim to enhance the current stratification approach by incorporating archetypical characteristics that drive malaria, including environmental, access, epidemiological, and socioeconomic factors. An archetype-based stratification approach may inform the tailoring of interventions to accelerate progress toward elimination. We used literature-informed data from 2017 to 2024 from the Zambia Health Information Management System and publicly available datasets. A zero-inflated negative binomial mixed-effects regression model was fitted to identify significant variables influencing malaria heterogeneity at the HFCA level. The significant variables were used to archetype HFCAs using the Clustering Large Applications (CLARA) algorithm. Significant variables included: walking travel time to the nearest health facility; percentage of malaria attributed to pregnant women; percentage of malaria attributed to children under five (U5); Children U5’s bednet (Long-lasting insecticide-treated nets or Insecticide-treated nets) coverage; bednet use rate; elevation; enhanced vegetation index; precipitation; and housing quality. Four new archetypes were derived. Relative to malaria incidence-based stratification, the archetype-based stratification exhibited spatiotemporal stability over time. Across all four new archetypes, elevation, bednet use rate, and housing quality emerged as the most influential features. However, all four archetypes exhibited distinct features influencing their makeup. The study proposes embedding archetypical characteristics in the malaria incidence-based stratification approach to better inform intervention tailoring and prioritisation based on the distinct archetype features driving malaria at the HFCAs level. Furthermore, the study provides a precursor for simulating potential intervention mixes based on malaria transmission archetypical characteristics, which can provide decision-makers with projected impact of varied investments in HFCAs with similar archetypical features.

## Introduction

Similar to many infectious diseases, malaria is characterised by clustering tendencies, perpetuated by the heterogeneity in various factors that provide a suitable environment for mosquitoes to thrive and parasite transmission [1]. The clustering trend is observed at the global level, where more than 90% of the more than 200 million global malaria cases have been historically attributed to sub-Saharan Africa [2,3]. The clustering tendency is also observed at national scales, including Zambia [1]. Like many sub-Saharan countries, Zambia is also burdened with malaria, with approximately eleven million cases reported at health facilities and community structures, and contributing about 1.4% to the global malaria burden as of 2023 [2,4,5]. Despite Zambia not being among the highest contributors to global malaria cases, its malaria risk distribution (annual cases per 1,000 population) and malaria parasite prevalence are characterised by spatial heterogeneity over smaller spatial boundaries, such as health facility catchment areas (HFCAs) [6].

The underlying spatial differences in the malaria burden in Zambia are driven by variations in behavioural, access, environmental, ecological, socioeconomic, demographic, and malaria intervention histories [6–8]. Furthermore, the Zambia National Malaria Elimination Centre (NMEC) identified rural populations, people living at lower altitudes and near water bodies, individuals with low socioeconomic status, mobile populations, pregnant women, and children under five (U5) as vulnerable populations for malaria infection [6].

Over the years, the government of Zambia and several partnering organisations have delivered various malaria intervention strategies [6,9]. Building on past experiences and the World Health Organisation’s (WHO) recommendations, Zambia’s 2022-2026 National Malaria Elimination Strategic Plan (NMESP) prescribed intervention packages for HFCAs, based on their malaria incidence profiles, which is a stratification of annual cases per 1,000 population [6,9–11]. For example, HFCAs with greater than 500 annual cases per 1,000 people pursue intensified control strategies (reducing transmission), while HFCAs with fewer than 50 cases pursue pre-elimination strategies (interrupting local transmission) [6]. However, several interventions function as control strategies or pre-elimination strategies [6,12]. Considering the widening shortfalls in funding for malaria, not all interventions per strategy can be deployed [11,13]. As such, it is important to determine which intervention to implement and where to gain the most from these interventions [8,14,15].

In the call for optimal allocation of interventions, the WHO emphasises doing away with a one-size-fits-all approach and rather applying interventions that are tailored to local settings [12]. This is because the effectiveness of interventions is highly influenced by how well they align with the unique contextual realities of specific areas [16–19]. To align with the global emphasis of deploying optimal interventions and to contribute towards the global malaria goals, the Zambian government and partnering organisations have been taking deliberate actions to pilot and deploy targeted interventions in selected parts of the country [6,17,20]. However, other than the HFCA malaria incidence profiling, no country-wide stratification has been conducted to inform the tailoring of interventions at a national level based on malaria drivers or factors that may influence intervention effectiveness [6].

The tailoring of interventions beyond malaria incidence profiles requires incorporating factors that drive transmission [18]. The incorporation of malaria drivers in the stratification process not only provides a relatively robust framework for intervention tailoring but also identifies opportunities for de-prioritisation and reallocation of resources [21]. Due to the clustering tendency of malaria, several studies, including Ozodiegwu et al. (2023) for Nigeria, have used cluster analysis to generate archetypical malaria profiles using malaria drivers as part of their stratification process[15].

To enhance Zambia’s intervention targeting approaches, this study leverages and builds on the existing stratification strategy, which allocates interventions based on HFCA-level malaria incidence profiles. The study achieves this by identifying key malaria drivers at the HFCA level and uses them to cluster HFCAs into new archetypes with similar characteristics. It then integrates the archetypes with the malaria incidence levels prescribed by the NMSP. Furthermore, the study examines the spatio-temporal stability of the archetypes compared to the malaria incidence based HFCA stratification. To inform intervention tailoring, the study also highlights the most influential drivers that determine the clustering tendency per individual archetype.

## Methods

### Data

Yearly HFCA level data from 2017 to 2024, sourced from the Zambia Health Analytics Platform (ZHAP), along with publicly available geospatial data, were used in this study. The platform aggregated data from the Zambia Health Management Information System (HMIS) and other sources. Geospatial variables such as walking time to health facilities (S1 Table provides a comprehensive list, descriptions, and sources of all variables explored) were extracted from raster files using the terra package [22] using the HFCA boundaries shapefile. The HFCA boundaries shapefile was obtained from the NMEC and was generated in 2023. To maintain the geospatial capability of the analysis, the HFCA boundaries were used as the master source for merging HFCA data. Out of the 3285 unique facilities from 2017 to 2024 in ZHAP, 563 were not available in the HFCA shapefile dataset, which contained 2722 HFCA boundaries. As a result, the 563 facilities were excluded from the analysis. The excluded facilities were either: health posts that were not allocated their own HFCA boundaries and were located within the HFCA boundaries of the main health facility; facilities that had been split before or after the generation of the HFCA shapefile boundaries; or hospitals that do not have catchment areas as they function as referral facilities.

### Descriptive Analysis

To understand the distribution of malaria burden at the HFCA level, we categorised annual malaria cases using the NMEC stratification: ‘no malaria’ (0 cases), ‘very low’ (1–49), ‘low’ (50–199), ‘moderate’ (200– 499), and ‘high’ (>500) cases per 1,000 population per year. However, we grouped “no malaria” (0 cases) and “very low” (between 1 and 49 cases per 1,000 population per year) into a single category labelled “very low” (0–49) because there were very few “no malaria” HFCAs [6]. These categories were then mapped to visualise changes in malaria incidence stratification across HFCAs nationwide from 2017 to 2024, along with a trend bar chart.

### Covariate Selection for Cluster Analysis

To select variables for cluster analysis, the study needed to reduce the number of variables to the ones driving malaria incidence at the HFCA level. As such, available literature-informed malaria drivers and intervention data in S1 Table were examined for potential inclusion in the cluster analysis. The study explored three variable reduction methods, including Principal Component Analysis (PCA), Singular Value Decomposition (SVD), and regression. The regression method consisted of two steps: a Pearson correlation coefficient test to identify and eliminate correlated variables, and a zero-inflated negative binomial mixed-effects regression model to determine which variables were significantly associated with malaria incidence. From the Pearson correlation coefficient test, variables that were not correlated with others were considered for the regression. For pairs of variables with a correlation coefficient greater than 0.6, only the variable with the stronger association with malaria was retained, while the other was excluded [23]. The Pearson correlation coefficient test approach prevented the subsequent statistical model from multicollinearity and informed the elimination of relatively unimportant variables [23].

A zero-inflated negative binomial mixed effects regression model, with the annual number of malaria cases in the HFCA as the dependent variable and the covariates identified in the Pearson correlation coefficient test process as regressors, was fit [24]. The model was fit using the glmmTMB package in R and included the HFCA unique identification number as a random effect nested at the province level [24]. The province nesting was included to account for varying provincial dynamics, such as political and donor presence.

### Clustering Approach

The HFCA level PCA and SVD reduced, and significant covariates in the regression model were each used to classify HFCAs. The study explored hierarchical clustering, K-means, K-Medoids Clustering (PAM), and Clustering Large Applications (CLARA) algorithms using data from the three data reduction methods. The process involved scaling the variables to have a mean of zero and a standard deviation of one, using the scale function from the base package in R. Thereafter, the optimal number of clusters was determined for each variable reduction methods and clustering algorithm using the Ward’s method for hierarchical clustering and the silhouette and within-cluster sum of square (wss) methods for K-means, PAM, and CLARA algorithms. To select the appropriate data reduction method, clustering algorithm, and the optimal number of clusters, the study evaluated the clustering quality for each combination. Clustering quality was evaluated using the Silhouette Score displayed in Equation 1 [25–27], which quantifies how well each cluster point fits within its assigned cluster compared to its proximity to points in other clusters. Having a scale ranging from -1 to 1, a higher Silhouette Score indicates that clusters are clearly distinguished and apart [25–27].

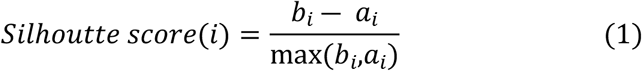

Where,

- *a*_*i*_ is the smallest mean distance of *i* to points in the cluster to which it was assigned [25,26].
- *b*_*i*_ is the smallest mean distance of *i* to the points in the nearest cluster to which it was not assigned [25,26].

After selecting the clustering algorithm and the final number of clusters with the highest score, the clustering results were mapped temporally. Since the data used for this analysis is longitudinal, some HFCAs had missing data due to the splitting and formation of new catchments over time. In these cases, the study assigned such HFCAs the cluster of their nearest neighbour as their archetype.

### Cluster Feature Importance and Sensitivity Analysis

To measure the importance of each feature, the percentage change in the silhouette score was recorded when each feature was removed from the analysis. Similarly, the individual cluster-level silhouette score percentage change was also recorded to capture the influence of each feature on individual clusters.

## Results

### Spatio-temporal Heterogeneity Characterises Zambia’s Malaria Incidence Stratification at the HFCA Level, Except For Southern Province

Malaria incidence stratifications, determined by the NMEC categorisation of cases per 1,000 population per year, varied heterogeneously at the HFCA level (Fig 1). In this figure, NA implies missing data due to various reasons, such as the splitting of HFCA. The malaria incidence trend observed in the data suggests that the southern province has historically maintained a very low incidence profile in most of its HFCAs in the analysis period (Fig 1). Relative to the very low-risk stratum, HFCAs in other categories transition back and forth across the incidence levels over the years (Fig 1). Additionally, as shown in Fig 1b, malaria incidence trends have stagnated, except for 2023, when the high-risk category (500+) increased to 47% from 38% in 2022. The stagnant trend observed in the period 2021-2024, as shown in the figure, is similar to that of 2017-2020, where incidence strata levels fluctuate slightly without a consistent reduction or increase.

**Fig 1.**
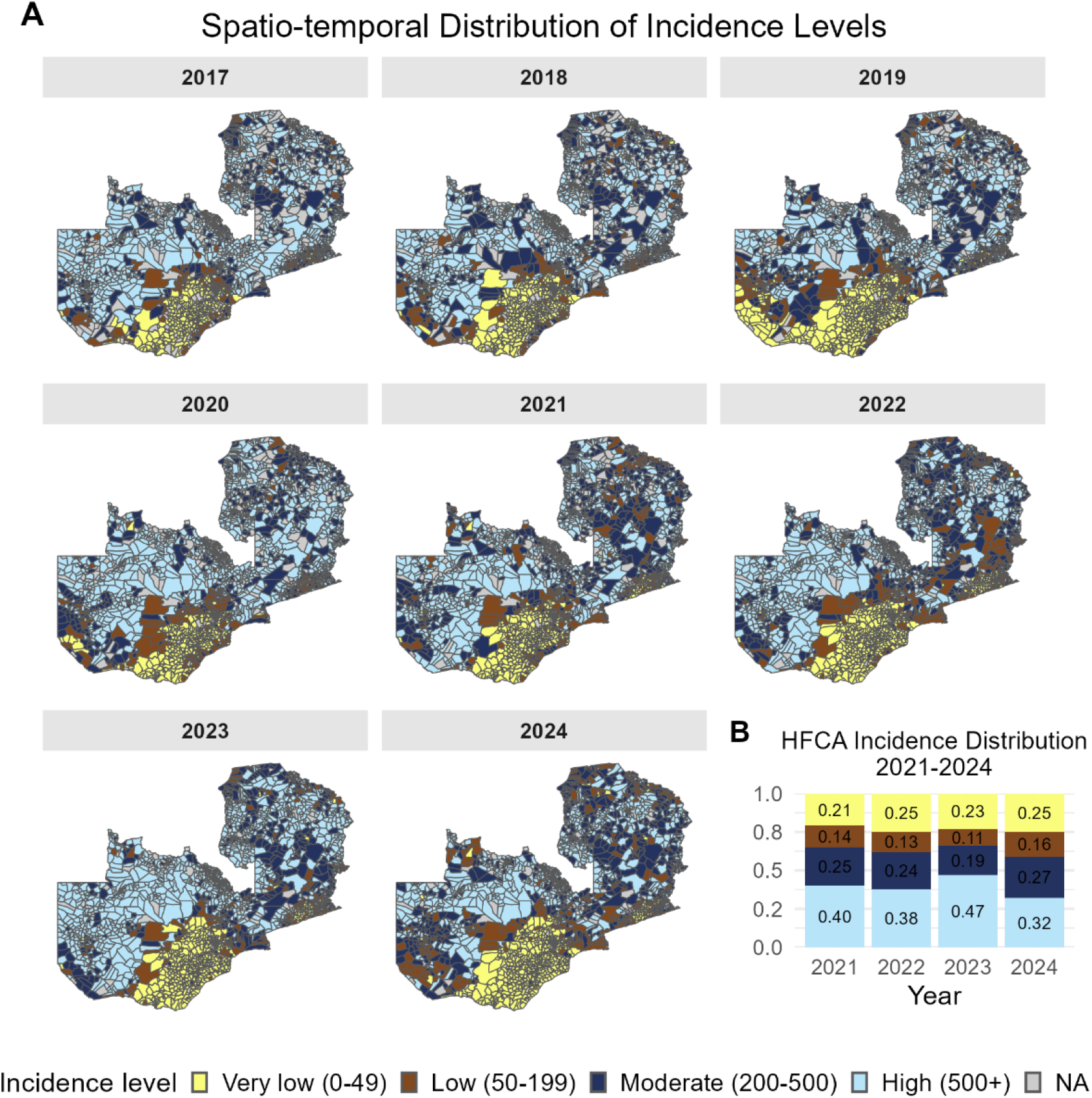
Spatio-temporal malaria incidence stratification distribution at the health facility catchment area (HFCA) level: (A) HFCAs mapped by year and categorised by malaria incidence stratification as defined by NMEC. In this figure, NA implies missing data (B) Percentage distribution of malaria incidence stratified as defined by NMEC from 2021 to 2024.

### Identified and Eliminated Variables for Cluster Analysis

Table 1 presents the results from a Zero-inflated negative binomial mixed-effect regression model, with malaria incidence per 1,000 population as the dependent variable. The independent variables of interest, shown in Table 1, except for “Year”, emerged as the preliminary variables of interest after the correlation coefficient test. Thus, they were either not correlated with other variables, or they were more correlated with malaria incidence relative to the variable(s) they were correlated with, which were eliminated. The model results suggest that we failed to find a significant association between the percentage of intermittent preventive treatment in pregnancy (at least three doses), indoor residual spraying coverage, and malaria incidence at the HFCA level. Therefore, the two variables were eliminated in the subsequent cluster analysis. Nonetheless, the model identified that walking travel time to health care, percentage of malaria attributed to pregnant women, percentage of malaria attributed to children (U5), children U5 bednet coverage, bednet use rate, elevation, enhanced vegetation index, precipitation, and housing quality are highly associated with malaria incidence.

**Table 1:** Zero-inflated negative binomial mixed-effect regression model estimates for malaria incidence at the HFCA level from 2017 to 2024.

| Variable | Estimate | Std. Error | z value | Pr(>\|z\|) |
| --- | --- | --- | --- | --- |
| Intercept | 3.69 | 0.451 | 8.189 | 0.000 |
| Year2018 | -0.125 | 0.021 | -5.917 | 0.000 |
| Year2019 | -0.153 | 0.023 | -6.551 | 0.000 |
| Year2020 | 0.098 | 0.02 | 4.991 | 0.000 |
| Year2021 | -0.145 | 0.022 | -6.746 | 0.000 |
| Year2022 | -0.274 | 0.021 | -12.791 | 0.000 |
| Year2023 | -0.043 | 0.022 | -1.911 | 0.056 |
| Year2024 | -0.344 | 0.027 | -12.683 | 0.000 |
| Walking travel time to health care in minutes | 0.519 | 0.157 | 3.314 | 0.001 |
| Percentage of malaria attributed to pregnant women | -8.197 | 0.243 | -33.709 | 0.000 |
| Percentage of malaria attributed to children (U5) | 0.316 | 0.109 | 2.891 | 0.004 |
| U5 children's bednet coverage | 0.195 | 0.025 | 7.652 | 0.000 |
| Percent of intermittent preventive treatment in pregnancy (at least 3 doses) | 0.005 | 0.022 | 0.223 | 0.824 |
| Bednet use rate | 2.04 | 0.222 | 9.183 | 0.000 |
| Indoor residual spraying coverage | -0.01 | 0.036 | -0.272 | 0.786 |
| Elevation | 0.01 | 0.001 | 11.02 | 0.000 |
| Enhanced vegetation index | 0.102 | 0.016 | 6.299 | 0.000 |
| Precipitation | 0.116 | 0.034 | 3.395 | 0.001 |
| Percentage of good-quality housing | -4.258 | 0.236 | -18.046 | 0.000 |

Trend-wise, the data suggest that incidence significantly differed from 2017 (the reference year) in all years except 2023. However, holding all other variables constant, the “year” variable estimates fluctuated over time, indicating the absence of a consistent trend, which statistically confirms the descriptive findings in Fig 1.

On the other hand, both PCA and SVD methods reduced the variables from 20 to 16 at a 95% threshold (S1 Fig), compared to the eight achieved by the regression approach.

### Archetype Identification and Validation

Several optimal numbers of clusters, ranging from three to seven, were identified by the Hierarchical, K-means, PAMS, and CLARA methods, using data reduced by PCA, SVD, and regression. Among the four clustering methods, CLARA’s four-cluster option using regression-reduced data yielded the highest average silhouette score of 0.25, emerging as the best clustering option for defining the archetypes. In contrast, the silhouette scores for both SVD and PCA were relatively lower than those using the eight variables, despite using the same clustering algorithms and number of clusters. As such, the regression approach was selected as the appropriate covariate reduction method.

### Identified Archetypes are Relatively Spatially Homogeneous Across HFCAs Compared to Incidence Stratification

Fig 2 shows the spatial distribution of archetypes that each HFCAs was categorised by, derived through cluster analysis from 2017 to 2024. Based on the prevailing malaria drivers in a particular year, some HFCAs transitioned into different archetypes. Relative to the incidence-based stratification shown in Fig 1, the identified archetypes, as shown in Fig 2, exhibit geographic continuity across all years, especially the most recent years. Over the years, we observed that the majority, 38-47%, of HFCAs belonged to archetype B (Fig 2b). We also noted that approximately one percent of the HFCAs belong to the third archetype, which visually coincides with Zambia’s major “line of rail”, a geographical landmark that defines the country’s relatively developed stretch (Fig 2) [28].

**Fig 2.**
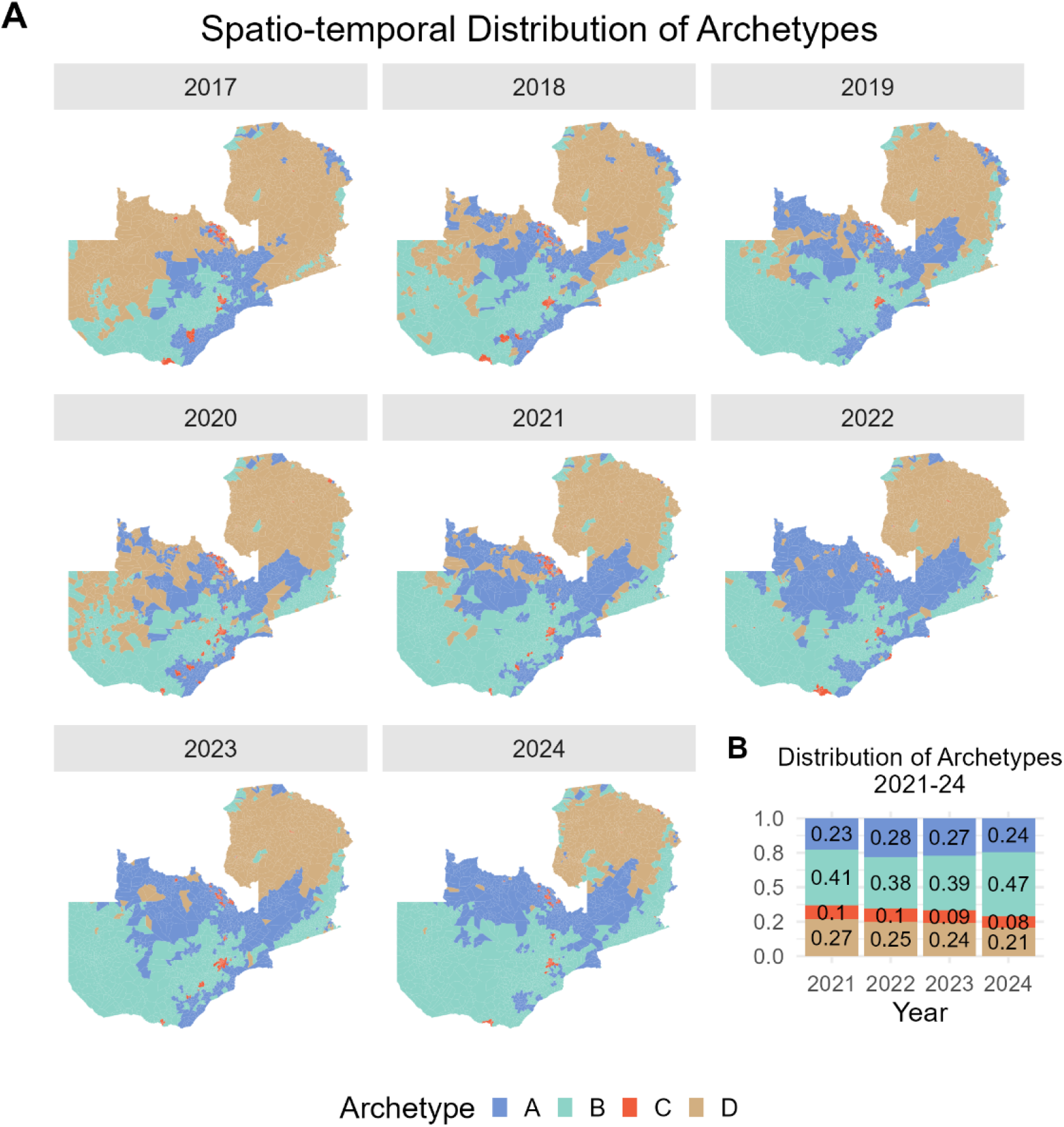
Spatio-temporal distribution of archetypes at the HFCA) level: (A) HFCAs mapped by year and categorised by archetype. (B) Archetype percentage distribution from 2021 to 2024 at HFCA level.

### Identified Archetypes Exhibit Relatively More Spatio-temporal Stability Than Incidence-based Stratification

Figs 3a and 3b show Sankey diagrams of HFCA malaria incidence stratification and archetypes, respectively, from 2017 to 2024. In these figures, nodes (vertical bars) represent values in the respective categories for each year, and arcs represent transitions connecting nodes across years, illustrating the HFCAs’ movement from a source year to a target year. Furthermore, the size of the arcs represents the number of HFCAs that transition from the source node to the target (Figs 3a and 3b). Therefore, the thicker the arc, the larger the number of HFCAs.

**Fig 3.**
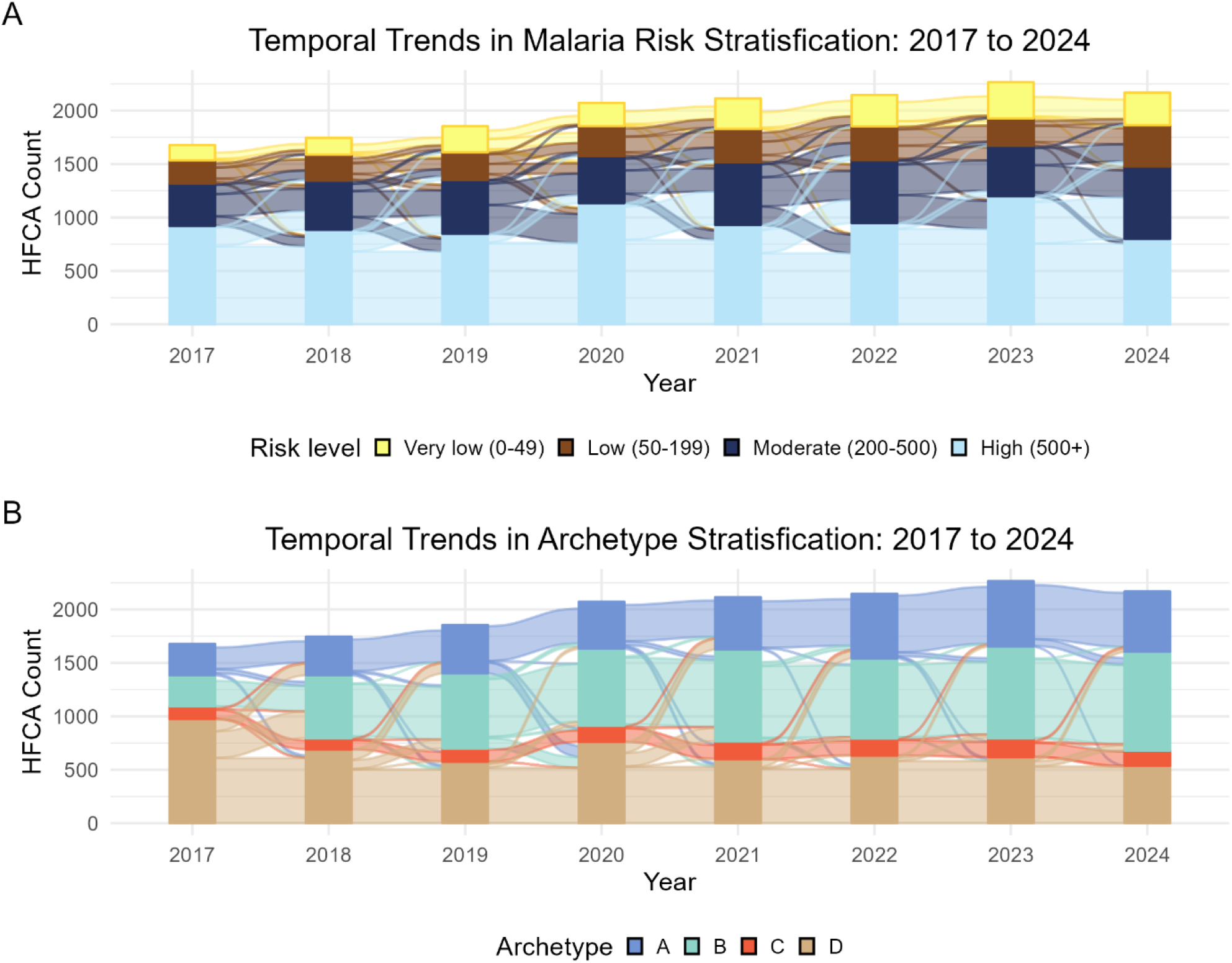
Temporal stability of health facility catchment classifications:(A) Incidence-based stratification trends from 2021 to 2024. (B) Archetype-based classification trends from 2021 to 2024.

From 2017 to 2024, there has been an upward trend in the number of HFCAs, as some HFCAs were divided to allow the birth of new facilities. Relative to Fig 3b, the arcs in Fig 3a are thicker, suggesting more facility transitions between incidence levels over time than archetypes. This suggests that archetypes are relatively more stable over time compared to incidence categorisation. Notably, we see that most of the facilities transition from the moderate (200 – 499) incidence level to other levels (Fig 3a). Additionally, very few HFCAs transition into and out of the “very low (0-49) category (Fig 3a). Similarly, almost none of the HFCAs transition from Archetype C.

### Malaria Incidence Stratification Distribution by Archetype and Overall Feature Importance

Distribution-wise, all archetypes comprised HFCAs from all incidence categories (Fig 4a). However, the proportion of very low transmission HFCAs was relatively small (only 0.4 percent) in Archetype D compared to the other archetypes (Fig 4a). This implies that 16 distinct HFCA characteristics strata emerge if the identified archetypes are disaggregated by malaria incidence stratification (Fig 4a). For instance, 54.9% of the HFCAs belong to Archetype A and the High-risk (500+) malaria incidence HFCA (Fig 4a).

**Fig 4:**
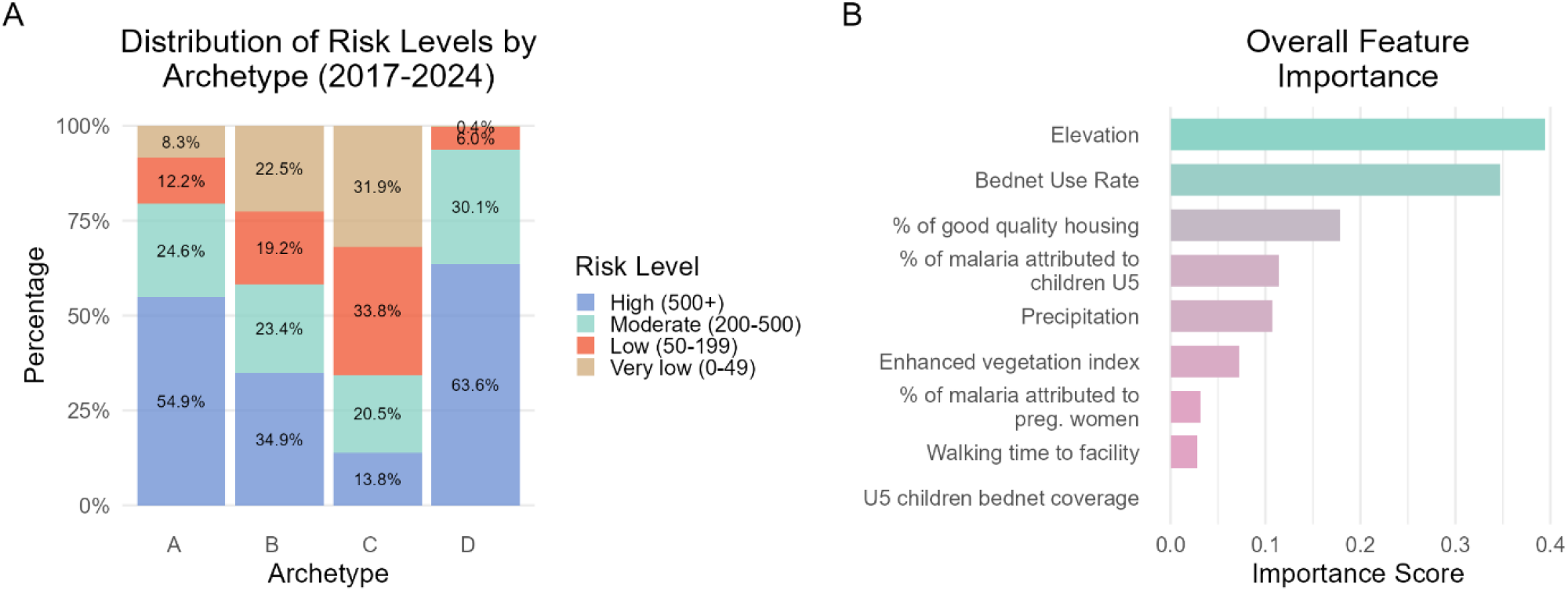
Archetype distribution among malaria incidence strata and feature importance: (A) Malaria incidence strata distribution per archetype for the analysis period 2017 to 2024. (B) Overall feature importance ranking in determining archetypes.

Fig 4b shows the overall importance ranking of features influencing HFCA clustering. The Y-axis represents the features, while the X-axis indicates the percentage influence each feature has on the overall clustering outcomes. As illustrated, elevation contributes approximately 39% to the clustering results (Fig 4b). Closely following elevation is the bednet use rate at around 35%, followed by the percentage of good housing quality (Fig 4b). Among all the features, bednet coverage for under-five children had the least influence of approximately 2% (Fig 4b).

### Distribution of Malaria Drivers and Feature Sensitivity by Archetypes

Fig 5 summarises the distribution of variables across archetypes to characterise their composition. In Fig 5, all variables were scaled to fall between zero and one. The influence of each variable was also measured by its sensitivity in determining the individual archetypes, as shown in Table 2 and S2 Fig.

**Table 2:** Summary of archetype features driving malaria and influencing individual archetypes

| Archetype | Features driving malaria relative to other archetypes | Influential features that also drive malaria |
| --- | --- | --- |
| **A** | • Low housing quality<br>• Low bednet use rate<br>• High EVI<br>• Long walking time to the nearest health facility<br>• Moderate precipitation | • Bednet use rate<br>• Walking time to the nearest health facility<br>• Precipitation |
| **B** | • Low housing quality<br>• Low elevation<br>• Long walking time to the nearest health facility | • Elevation<br>• Bednet use rate |
| **C** | • Moderate precipitation | • Precipitation |
| **D** | • Low housing quality<br>• High precipitation<br>• Low elevation<br>• Long walking time to the nearest health facility | • Precipitation<br>• Elevation |

**Fig 5.**
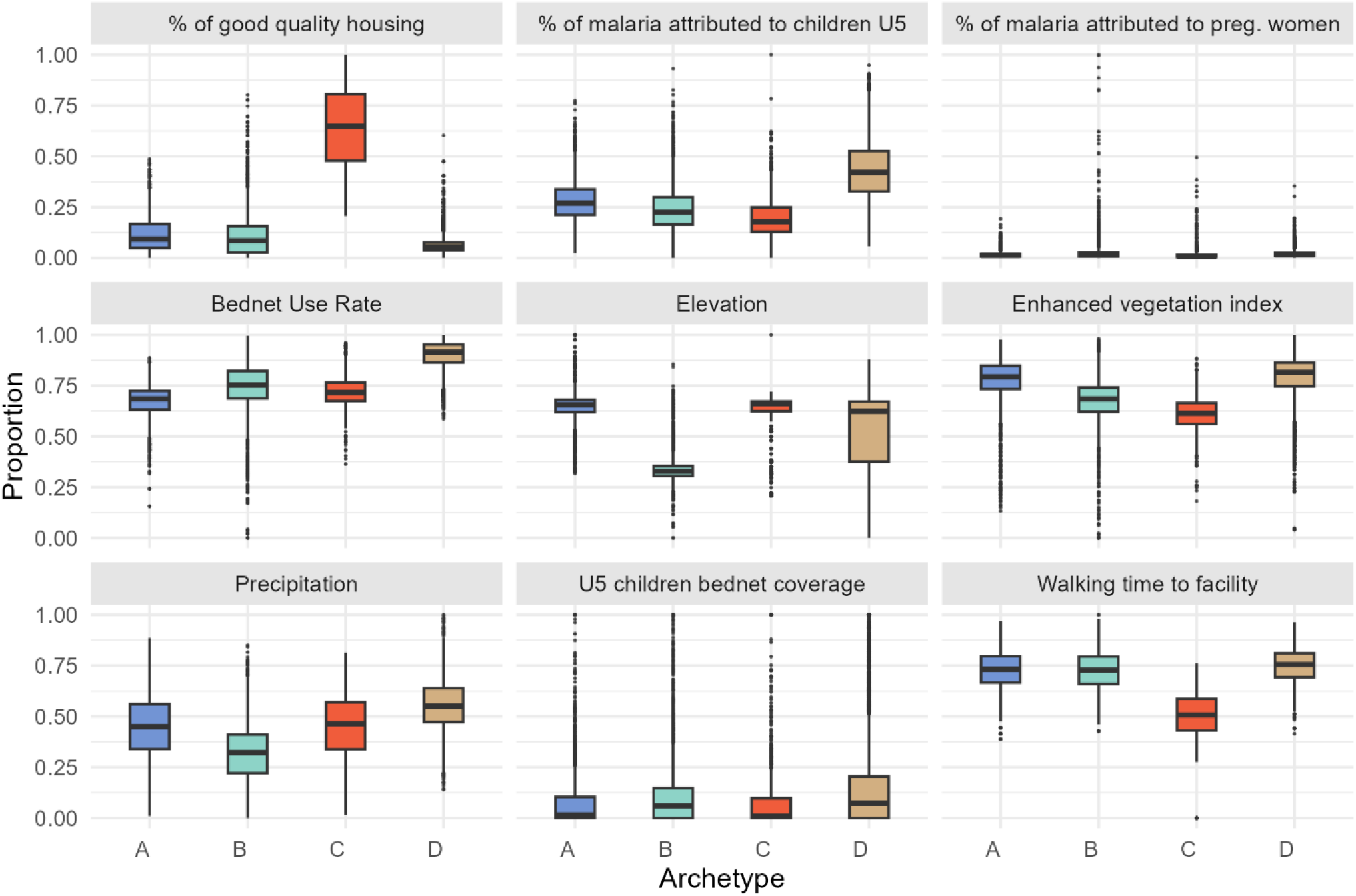
Distribution of normalised malaria drivers stratified by archetypes.

In terms of driving malaria cases, Fig 5 shows that Archetype A is characterised by relatively low housing quality, low bednet use rate, and long walking time to a health facility. However, only the bednet use rate and walking time to a health facility are among its most influential features (Table 2, and S2 Fig). Malaria-driving characteristics in Archetype B include relatively low housing quality, low elevation, and long walking distances to the nearest health facility (Fig 5). However, only elevation and bednet use rate are among its most influential features (Table 2, and S2 Fig). Archetype C is characterised by relatively low levels of all malaria drivers except for precipitation, which is also one of its most influential features (Table 2, and S2 Fig). Notably, in addition to coinciding with the “line of rail”, Archetype C is also characterised and influenced by high-quality housing. On the other hand, Archetype D is characterised by relatively low-quality housing, high precipitation, low elevation, and long walking time to the nearest health facility (Fig 5). Among these, precipitation and elevation are its most influential malaria drivers (Table 2, and S2 Fig).

## Discussion

We analysed HFCA level data for Zambia. Our objectives were to identify variables influencing malaria heterogeneity at the HFCA level to cluster the HFCAs into archetypes with similar characteristics, and to compare the spatio-temporal stability between malaria incidence and archetype stratification approaches. These analyses were undertaken to build on the existing stratification approach and to inform the tailoring of malaria interventions at the HFCA level.

The analysis of malaria incidence stratification showed a non-constant trend from 2017 to 2024. Malaria spatial heterogeneity was observed nationwide at the HFCA level, except in the southern province of the country, where the majority of HFCAs had very low incidence (0-49). The very-low incidence observed in the majority of Southern province HFCAs is likely due to the deliberate scale-up of malaria control and targeted elimination efforts in the province since the 2010s [29]. To explain the heterogeneity of malaria incidence across HFCAs, eight variables were identified in the pre-clustering variable selection process. Interestingly, these variables represented most of the thematic areas known to influence malaria transmission, as well as the vulnerable populations identified by the NMEC [6]. The variables included: walking travel time to healthcare representing access; the percentage of malaria attributed to pregnant women and children U5 representing the vulnerable population; children U5 bednet coverage, intermittent preventive treatment in pregnancy (at least three doses), and IRS coverage reflecting intervention history; bednet use rate capturing behavioural factors; elevation, EVI, and precipitation representing environmental factors; and the percentage of good-quality housing capturing socioeconomic conditions. However, intermittent preventive treatment in pregnancy (at least three doses) and IRS coverage were not statistically significant.

Relative to malaria incidence stratification, the established clusters from the identified variables exhibit geographic continuity across all years, particularly in the most recent ones, which represents a tendency of how factors such as precipitation and elevation manifest geographically [18]. Furthermore, the locations of archetypes, such as Archetype D, which is influenced by precipitation, coincided with regions (northern) known to have the highest levels of rainfall [30]. Archetype C, which is highly influenced by good-quality housing, coincided with Zambia’s “line of rail,” a landmark used to describe relatively developed areas of the country [28]. The correspondence between the established archetypes and national attributes further supports the validity of our clustering approach.

Except for HFCAs with very low malaria cases (0-49), a relatively higher number of HFCAs switched from one malaria incidence category to another over time, while fewer HFCAs changed their archetype. The shifts in malaria incidence classification among HFCAs may be attributed to several factors, including the adjustments to intervention strategies [6,31,32]. Typically, HFCAs with higher case counts receive more intensive malaria control interventions [6]. When case counts decline, these areas may transition to pre-elimination strategies [6]. However, if the new, less intensive interventions are not well matched to local conditions, or if neighbouring areas continue to have high transmission, it may lead to a resurgence in cases and a return to relatively higher-incidence strata [6,31–33]. In contrast, the archetype-based approach offers more stability, which may inform package switching to consider the archetypical characteristics of the HFCAs, which may aid consistent progress in malaria reduction, as the interventions would be better tailored to the underlying context and needs of each HFCA [15,18,34]. Moreover, relying solely on malaria cases is prone to spatial error due to various reasons, such as the misclassification of patients from different catchments who prefer certain health facilities over those in their catchments [17]. The implication of such situations may result in deploying intervention packages in areas where cases are recorded, rather than in areas where they originate [17].

The distinct features characterising the established archetypes can be used to tailor interventions aimed at reducing malaria incidence. These include low bednet use and long walking time to a health facility for Archetype A; low elevation and low bednet use for Archetype B; moderate precipitation for Archetype C; and low elevation and high precipitation for Archetype D. For Archetype A, appropriate interventions would include strengthening social and behaviour change communication to increase bednet use, alongside improving healthcare access through strategies such as the establishment of more health posts, and/or mass testing and treatment [7]. For Archetypes C and D, further exploration of precipitation seasonality may be necessary to ensure appropriate timing of interventions [15,18]. Interestingly, Archetype C has relatively few malaria-driving features influencing it. As such, HFCAs within Archetype C could be deprioritised in terms of interventions, particularly those targeting the socioeconomic determinants of malaria, since it is mainly influenced by good housing quality, which may be a proxy for high socioeconomic status, while the rest of the malaria influencing features are relatively low [21,35,36].

In the context of building on the current stratification approach, interventions within the control and pre-elimination packages may be prioritised based on the archetypical makeup of HFCA. This can be achieved by identifying the HFCA intersection between malaria incidence strata and archetypes, as illustrated in S1 Text, to inform intervention allocation and prioritisation from the prescribed malaria incidence stratification packages. For example, interventions such as reactive case detection that are typically implemented in HFCA belonging to the very-low (0 - 49) malaria incidence stratum can be prioritised in HFCAs that belong to both the very-low (0 - 49) malaria incidence category and Archetype A where access to healthcare is limited due to long walking times to health facilities [6]. Since the data used and archetypes generated are longitudinal, an averaged dataset using the same algorithm and number of clusters can be used to inform the enhancement of the malaria incidence stratification, as shown in S1 Text. This implies that 16 distinct characteristics of HFCAs would emerge to guide the package allocation in the proposed stratification, based on the four malaria incidence stratification profiles multiplied by the four newly identified archetypes.

It is also worth noting that the study provides a precursor for more formal investigations to assess the impact of various interventions for the identified archetypes through relatively sophisticated approaches, such as mathematical models. Considering the challenges associated with historical malaria data, including issues of quality and availability at finer spatial scales [37,38]. Archetypical characteristics also offer a valuable basis for projecting the outcomes of specific interventions from one HFCA to another [18]. These projections can be derived from modelling outcomes of HFCAs within the same archetype that have good-quality data available [18].

While this study provides valuable insights into archetypical makeup of HFCAs, we acknowledge certain limitations. The clustering quality, as indicated by a silhouette score of 0.25, reflects the complexity of working with high-dimensional data, which can affect clustering performance [39]. This effect was particularly evident when using relatively higher-dimensional datasets (16 features) derived from PCA and SVD, which resulted in even lower validation scores (approximately 0.15 to 0.18). Despite these challenges, the current approach strikes a practical balance between clustering quality and the inclusion of a sufficient number of archetype features to meaningfully inform appropriate intervention allocation. It is also worth noting that some variables in our study were derived from modelled estimates, which may introduce a degree of uncertainty into the outcomes.

Given the multifaceted nature of malaria, this work serves as an important starting point for further refinement of archetype-based approaches. Future research could explore incorporating additional factors such as seasonality and health system characteristics, which may improve clustering outcomes. To leverage the benefits of interventions tailored to archetypes, our future work includes mathematical modelling to assess how intervention efficacy varies across different archetypes to inform the optimisation of intervention mixes. Additionally, further research is warranted to better understand why some HFCAs transition between malaria incidence levels over time.

## Conclusion

The study identified variables associated with malaria incidence at the HFCA level and used them to cluster HFCAs into archetypes with similar characteristics, to build on the existing stratification approach. The study established that incidence-based malaria incidence stratification is unstable over time compared to archetypical characteristics. As such, the study suggests embedding the archetypical characteristics in the stratification approach to better inform intervention tailoring and prioritisation. Furthermore, the study provides a precursor for simulating potential intervention mixes based on malaria transmission archetypical characteristics, which can provide decision-makers with the anticipated results of varied investments in HFCAs with similar archetypical features.

## Acknowledgements

The authors acknowledge the National Malaria Control Centre and the Modelling and Simulation Hub, Africa team, for their contribution to the ideas, feedback and code review.

## Author Contributions

**Chilochibi Chiziba:** Conceptualisation, Formal analysis, Investigation, Methodology, Software, Visualisation, Writing – original draft.

**Japhet Chiwaula:** Supervision, Validation, Writing – review & editing, Data Curation.

**Busiku Hamainza:** Supervision, Validation, Writing – review & editing.

**Sampa Chitambala-Otiono:** Supervision, Validation, Writing – review & editing.

**Sheetal Silal:** Conceptualisation, Funding acquisition, Methodology, Resources, Supervision, Validation, Writing – review & editing.

## Data Availability Statement

Malaria incidence and intervention data are available upon request from the National Malaria Elimination Centre. Sources for public data used in the study are highlighted in the SI Table.

## Ethics Approval Statement

This study was reviewed and deemed exempt for lack of human subjects by the Faculty of Health Sciences Human Research Ethics Committee (HREC) at the University of Cape Town in South Africa (HREC REF Number 178/2024) and the ERES Converge IRB in Zambia (2024-Jan-002).

